# Prediction of clinically relevant postoperative pancreatic fistula using radiomic features and preoperative data

**DOI:** 10.1101/2022.10.22.22281403

**Authors:** Nithya Bhasker, Fiona R. Kolbinger, Nadiia Skorobohach, Alex Zwanenburg, Steffen Löck, Jürgen Weitz, Ralf-Thorsten Hoffmann, Marius Distler, Stefanie Speidel, Stefan Leger, Jens-Peter Kühn

## Abstract

Clinically relevant postoperative pancreatic fistula (CR-POPF) can significantly affect the treatment course and outcome in pancreatic cancer patients. Preoperative prediction of CR-POPF can aid the surgical decision-making process and lead to better perioperative management of patients. In this retrospective study of 108 pancreatic head resection patients, we present risk models for the prediction of CR-POPF that use combinations of preoperative computed tomography (CT)-based radiomic features, mesh-based volumes of annotated intra- and peripancreatic structures and preoperative clinical data. The risk signatures were evaluated and analysed in detail by visualising feature expression maps and by comparing significant features to the established CR-POPF risk measures. Out of the risk models that were developed in this study, the combined radiomic and clinical signature performed best with an average area under receiver operating characteristic curve (AUC) of 0.86 and a balanced accuracy score of 0.76 on validation data. The following pre-operative features showed significant correlation with outcome in this signature (*p*<0.05)- texture and morphology of the healthy pancreatic segment, intensity volume histogram-based feature of the pancreatic duct segment, morphology of the combined segment, and BMI. The predictions of this pre-operative signature showed strong correlation (Spearman correlation co-efficient, *ρ* = 0.7) with the intraoperative updated alternative fistula risk score (ua-FRS), which is the clinical gold standard for intraoperative CR-POPF risk stratification. These results indicate that the proposed combined radiomic and clinical signature developed solely based on preoperatively available clinical and routine imaging data can perform on par with the current state-of-the-art intraoperative models for CR-POPF risk stratification.

## Introduction

Despite advances in conservative and surgical treatment approaches, pancreatic cancer remains one of the deadliest malignant diseases. All curative treatment approaches necessitate surgical resection. However, 80-85% of patients are diagnosed with locally advanced or metastasised (thus unresectable) tumours^1,2^. Surgical treatment options for patients with tumours of the pancreatic head include pylorus-resecting (Whipple surgery) and pylorus-preserving pancreaticoduodenectomy (PPPD). These surgical procedures are complex and are associated with severe postoperative complications like postoperative pancreatic fistula (POPF), which can significantly delay or impede adjuvant chemotherapy, thus affecting the oncological treatment course and outcome^3^. POPF is caused by pancreato-enteric anastomotic leakage^4^ and affects about 10-30% of patients undergoing pancreatic head resection^5–7^. The definition of POPF published in 2016 by the International Study Group on Pancreatic Fistula Definition (ISGPF) distinguishes between asymptomatically elevated levels of drain amylase (termed biochemical leak), and clinically relevant (CR-)POPF, which necessitates invasive treatment and leads to prolonged hospital stay^5,8^.

A plethora of risk stratification methods have been introduced in the past decade based on the association of CR-POPF with parenchymal risk factors like soft pancreatic parenchyma, small pancreatic duct width, and clinical factors like sex, intraoperative blood loss and body mass index (BMI)^6,9–13^. Most of these methods rely heavily on intraoperatively assessed factors like pancreatic duct width, haptically evaluated texture and blood loss for risk stratification. However, an early and accurate risk estimate based on the vast amount of available preoperative data has the potential to play a key role in pre-surgical decision-making, especially in case of high-risk cancer patients, where postoperative complications could delay or impede adjuvant therapy. For instance, small pancreatic duct size and large pancreatic remnant volume (PRV), estimated from preoperative computed tomography (CT) images, were found to be significantly associated with the risk of CR-POPF after pancreatic head resection^14–17^. A preoperative risk estimate could also complement intraoperative and postoperative findings for better perioperative monitoring.

It has been established that radiomic features (a large number of quantitative imaging descriptors extracted from medical images using data characterisation algorithms) contain good prognostic power in predictive modelling^18^. Skawran et al.^19^ investigated preoperative magnetic resonance imaging (MRI)-based radiomic features of the pancreatic body and tail in addition to pancreas-to-muscle T1 signal intensity ratio for CR-POPF prediction in a cohort of 62 patients undergoing pancreaticoduodenectomy and achieved an average area under the receiver operating characteristic curve (AUC) of 0.90 on the test data. Given that CT is the preoperative routine imaging modality in patients undergoing pancreatic resection, it would be preferable to predict the complication risk based on features extracted from CT imaging data rather than MRI data. A study conducted by Zhang et al.^20^ considered CT-based radiomic features from the pancreatic parenchyma remaining after tumour removal (healthy parenchyma) for postoperative prediction of CR-POPF, and achieved an AUC of 0.76 on the validation cohort. However, based on the above-mentioned clinically established risk factors such as small pancreatic duct size, one could deduce that considering pancreatic structures besides the healthy parenchyma for preoperative image based risk assessment might reveal interesting correlations with the surgical outcome.

The main goal of this study was to make use of preoperative data like CT-based radiomic features, mesh-based volumes of different pancreatic structures and clinical data to get an accurate and early risk estimate of CR-POPF that could complement the current intraoperative and postoperative risk stratification measures. The objectives of this study were (i) to develop risk models using CT-based radiomic features extracted not only from the normal pancreatic parenchyma but also from other intra- and peripancreatic anatomical structures, (ii) to compare the developed radiomic risk models with risk models that were developed using preoperative clinical data and mesh-based volumes of the selected anatomic structures, (iii) to quantitatively and qualitatively assess the feature importance for all the models using univariate analyses and visualisations, (iv) to analyse in detail the relationship between the relevant preoperative features and established risk factors for CR-POPF and, (v) to compare the predictions made by the proposed preoperative risk model with the intraoperative state-of-the-art risk score for CR-POPF risk prediction.

## Methods

### Patient characteristics

A total of 381 patients having undergone pancreatic surgery at the University Hospital Carl Gustav Carus Dresden between 2011 and 2019 were retrospectively screened for eligibility. Pancreatic head resection patients with a preoperative contrast-enhanced venous-phase CT containing a delineable pathology (tumour or cystic neoplasia) were selected for the study. The exclusion criteria were calcifying chronic pancreatitis or acute pancreatitis, presence of peripancreatic fluid collections, pancreatic pseudocysts, or a stent in the common bile duct. There were 108 patients after exclusion, all of whom had a clinical indication for surgical procedure and suffered from a localised pathology of the pancreas. The characteristics of this patient cohort are summarised in Table 1.

**Table 1.** Patient characteristics. Abbreviations: (CR-)POPF - (clinically relevant-) postoperative pancreatic fistula, BMI - Body mass index, CT - computed tomography, PPPD - pylorus-preserving pancreaticoduodenectomy, PDAC - pancreatic ductal adenocarcinoma.

| Characteristic | Statistic | CR-POPF | Non CR-POPF |
| --- | --- | --- | --- |
| Patients | n | 33 | 75 |
| Sex: | n |  |  |
| Male |  | 21 | 39 |
| Female |  | 12 | 36 |
| Age (in years) | mean ± standard deviation | 69.5 ± 8.8 | 68 ± 9.7 |
| BMI | mean ± standard deviation | 28.0 ± 5.2 | 24.0 ± 3.1 |
| Time delay between CT scan and surgery (in days) | mean ± standard deviation | 43.31 ± 47.49 | 75.12 ± 108.50 |
| Surgery type: | n |  |  |
| PPPD |  | 27 | 63 |
| Whipple surgery |  | 6 | 12 |
| Pancreatic pathology: | n |  |  |
| PDAC |  | 6 | 54 |
| Neuroendocrine tumour |  | 2 | 3 |
| Other tumour |  | 23 | 17 |
| Cystic neoplasia |  | 2 | 1 |

Frequency and severity of POPF were assessed for at least 30 days after the pancreatic head resection or until discharge from hospital, whichever occurred last. Following ISGPF standards, the CR-POPF endpoint included Grade B POPF (defined as a clinically evident fistula with mild restriction of patient condition, typically managed using antibiotic medication, transfusion, parenteral nutrition, prolonged drain maintenance or additional interventional drain placement) and Grade C POPF (a severe complication characterised by critical restriction of patient condition, typically necessitating intensive care or an operative intervention). Ethical approval for the retrospective analysis of clinical and imaging data was obtained from the Ethics Committee at the Technische Universität Dresden (BO-EK-263062020). All analyses were carried out in accordance with the relevant guidelines and regulations, in particular the Declaration of Helsinki and its later amendments. Written informed consent was waived by the Ethics Committee at the Technische Universität Dresden.

### Segmentation of the pancreas and image feature computation

For all patients, different anatomical structures within and adjacent to the pancreas were manually segmented in preoperative contrast-enhanced CT images (venous phase) using the picture archiving system PACS (Agfa Impax EE R20, Agfa Healthcare) and the open-source software Slicer 3D (version 4.10.2, www.slicer.org). In particular, the following structures were delineated (refer to Figure 1): pancreatic pathology (panc_pat), healthy pancreas (panc_heal), pancreatic duct (panc_duct), portal vein (pv), arteries (art), and bile duct (bile_duct). An additional segment was created by combining these delineated structures (combined). The annotation was performed by a radiologist (NS) with 5 years of experience in abdominal CT imaging, and was independently reviewed by a second radiologist (JPK) with 20 years of experience in pancreatic imaging. Any discrepancies between the two radiologists were resolved through discussion until a consensus was reached. The radiologists were blinded to the clinical data and outcomes of the patients during the annotation process. Subsequently, the imaging features were computed and extracted in compliance with the Image Biomarker Standardisation Initiative using the medical image radiomics processor (MIRP - www.github.com/oncoray/mirp)^21^.

**Figure 1.**
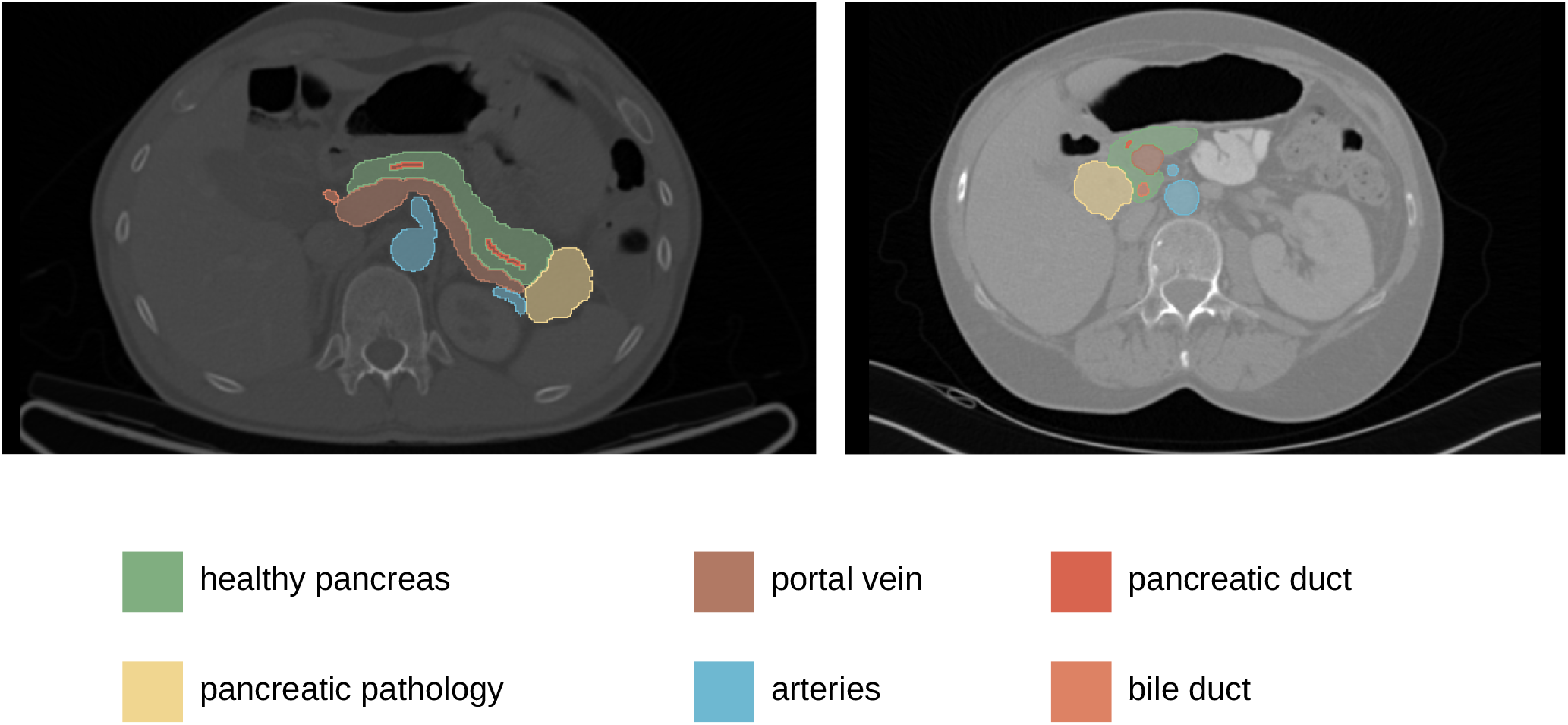
Examples of delineated structures: healthy pancreas, pancreatic pathology, portal vein, arteries, pancreatic duct, bile duct.

In brief, to correct differences in voxel spacing and slice thickness the CT images were resampled using linear image interpolation to a voxel size of 1.0 × 1.0 × 1.0 mm^3^. Subsequently, to emphasise image characteristics such as edges and blobs, nine additional images were created by applying spatial filtering to the base image. In particular, eight images were created using stationary coiflet-1 wavelet high-/low-pass filter along each of the three spatial directions and one image was created by applying a Laplacian of Gaussian (LoG) filter consisting of different filter kernel widths. Finally, for each segment statistical, histogram-based and texture features were calculated using the base and the nine transformed images. Furthermore, morphological features were computed for each of the pancreatic segments on the base image only. The configuration settings of the MIRP pipeline are summarised in Supplementary Table 2.

### Development of risk models

The familiar R package (version 0.0.0.54)^22^ was used for the development and validation of machine learning models to predict the risk of CR-POPF. The framework consisted of the following steps: feature pre-processing, feature selection, hyper-parameter optimisation, model development and model validation. All risk models were developed as previously described^23,24^ within a 10-fold cross validation scheme, repeated three times. Based on the entire retrospective cohort, the computed imaging features were transformed and normalised using Yeo-Johnson and z-normalisation methods, respectively. Subsequently, highly correlated features (absolute Spearman correlation coefficient, *ρ >* 0.9) were reduced using hierarchical clustering method. Feature selection was performed using 10 bootstrap samples (with replacement) of the training folds. The risk models were trained on these 10 bootstrap samples of the training folds, using selected features and optimised hyper-parameters^25^. Finally, an ensemble prediction was made by averaging the prediction scores of each model for the training and validation folds separately.

Different feature selection methods and learning algorithms were used for model development to reduce the risk of incidental findings^26^. In particular, the following four feature selection methods were used: mutual information maximisation (MIM), minimum redundancy maximum relevance (MRMR)^27^, lasso regression- and elastic net regression-based feature selection methods. For the learning algorithms, the following three approaches were considered: logistic regression without penalty, with lasso penalty and elastic-net penalty. The methods with lasso and elastic-net penalties prevent the models from overfitting by using a regularisation term^28^.

Various risk models were developed based on the radiomic features, the clinical variables, the mesh-based volumes of the annotated pancreatic segments and their combinations. An overview of the developed risk models is provided in Supplementary Table 6. The clinical variables used in the model development were: age, sex, BMI, preoperative presence of diabetes mellitus, smoking history, history of alcohol abuse, and preoperative lab values including CA 19-9 level and preoperative digestive enzyme levels (amylase and lipase). The continuous values for CA 19-9, amylase and lipase levels were transformed into binary values (0 - normal, 1 - abnormal) by considering their corresponding reference intervals used as a standard in the University Hospital Carl Gustav Carus Dresden. The normal reference intervals were: < 24 U/mL for CA 19-9, < 0.88 *μ*mol/s*L for amylase, and < 1 *μ*mol/s*L for lipase. The frequency, mean and standard deviation values of these variables for the used patient cohort are summarised in Table 1 and Supplementary Table 1.

A risk model developed using a selected subset of radiomic features is called a radiomic signature. The selection of a representative and stable radiomic feature subset is often a challenging task, especially when the number of features to be selected is significantly larger than the total number of samples^29^. This was true in our case, with 1706 radiomic features for each of the 108 patients. Therefore, we trained a set of preliminary risk models solely for the purpose of feature selection for the radiomic signature. The details of the preliminary risk model is included in the Supplementary section 3. Subsequently, representative features for pancreatic head resection patients were selected based on the permutation feature importance method. That is, features were deemed important and selected based on the reduction in model performance over the head resection patients when the most important features (based on feature occurrence) of the preliminary risk models were permuted. An ensemble risk model was eventually developed for only the head resection patients using the selected radiomic features. This model is henceforth referred to as the radiomic signature.

### Performance assessments

The performance of the models was assessed using the average area under the receiver operating characteristic curve (AUC) and balanced accuracy score on the ensemble predictions. The signatures were quantitatively and qualitatively examined to understand the decision rationale of various risk models. The significance of the association between individual features and CR-POPF outcome was assessed by performing univariate analyses using the logistic regression model with Benjamini-Hochberg adjustment for multiple testing^30^. The features with significant association with the CR-POPF outcome (*p <* 0.05) were standardised, and heatmaps of the resulting standardised features were visualised for the entire patient cohort in order to qualitatively assess the trends and correlations with the outcome. Additionally, the selected preoperative radiomic features were compared with established risk factors^16^ such as intraoperative pancreatic texture and pancreatic duct width, and preoperatively estimated pancreatic remnant volume (PRV). The standardised values of these established risk factors were plotted against the corresponding standardised values of the radiomic features. Moreover, CR-POPF risk predictions made by the model were compared to the intraoperatively assessed ua-FRS^13^.

## Results

Out of all the developed risk models, the model developed using a combination of the features selected for the radiomic signature and clinical variables, the combined radiomic and clinical signature, performed the best with an AUC of 0.86 and a balanced accuracy score of 0.76 on the validation data. Detailed results for all the models are included in the supplementary section 4. The feature expression map for the combined radiomic and clinical signature is illustrated in Figure 2. The features significantly associated (*p <* 0.05) with the outcome included two radiomic morphological features related to the panc_heal and combined segments, a radiomic texture feature related to the panc_heal segment, one radiomic intensity volume histogram (IVH) based feature related to the panc_duct segment, and BMI. Details about these radiomic features are mentioned in Supplementary Table 5.

**Figure 2.**
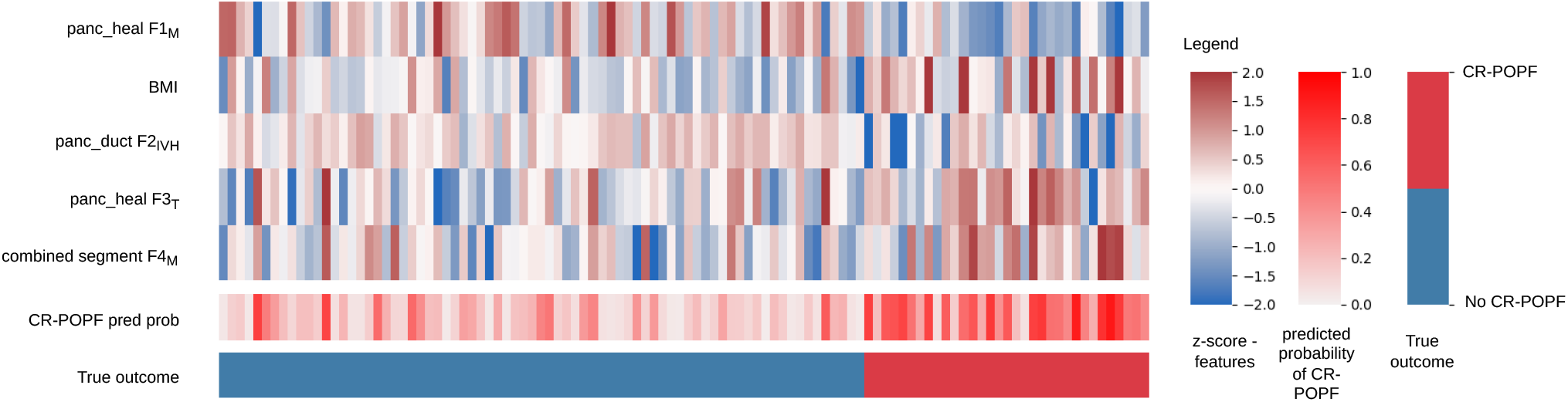
Feature expression maps for the combined radiomic and clinical signature. The values indicated in the heatmaps are z-scores of standardised features, probability of developing CR-POPF as predicted by the model and the true outcome for all the patients included in the study. Abbreviations: CR-POPF pred prob - predicted probability of CR-POPF, *F*_*M*_ - morphological feature, *F*_*T*_ - texture feature,, *F*_*IVH*_ - intensity volume histogram based feature.

To explore clinical interpretability of radiomic features, we evaluated the association between the features selected for radiomic signature and established risk factors for CR-POPF such as pancreatic texture, pancreatic duct width, and estimated PRV^16^ (Figures 3 and 4). Data for these established risk factors was available for 70 (out of 108) patients analysed in this study, out of whom 26 developed CR-POPF. We observed a moderate correlation of the estimated pancreatic remnant volume with the morphological feature *F*1_*M*_ of the healthy pancreatic segment (*ρ* = 0.61, Figure 3 a) and with the morphological feature *F*4_*M*_ of the combined segment (*ρ* = 0.54, Figure 3 b). We also observed a moderate association between the intensity volume histogram based feature *F*2_*IVH*_ of the pancreatic duct segment and the intraoperatively assessed pancreatic duct width (*ρ* = 0.53, Figure 4 a), and between the texture feature *F*3_*T*_ of the healthy pancreatic segment and intraoperatively determined pancreatic texture (Figure 4 b).

**Figure 3.**
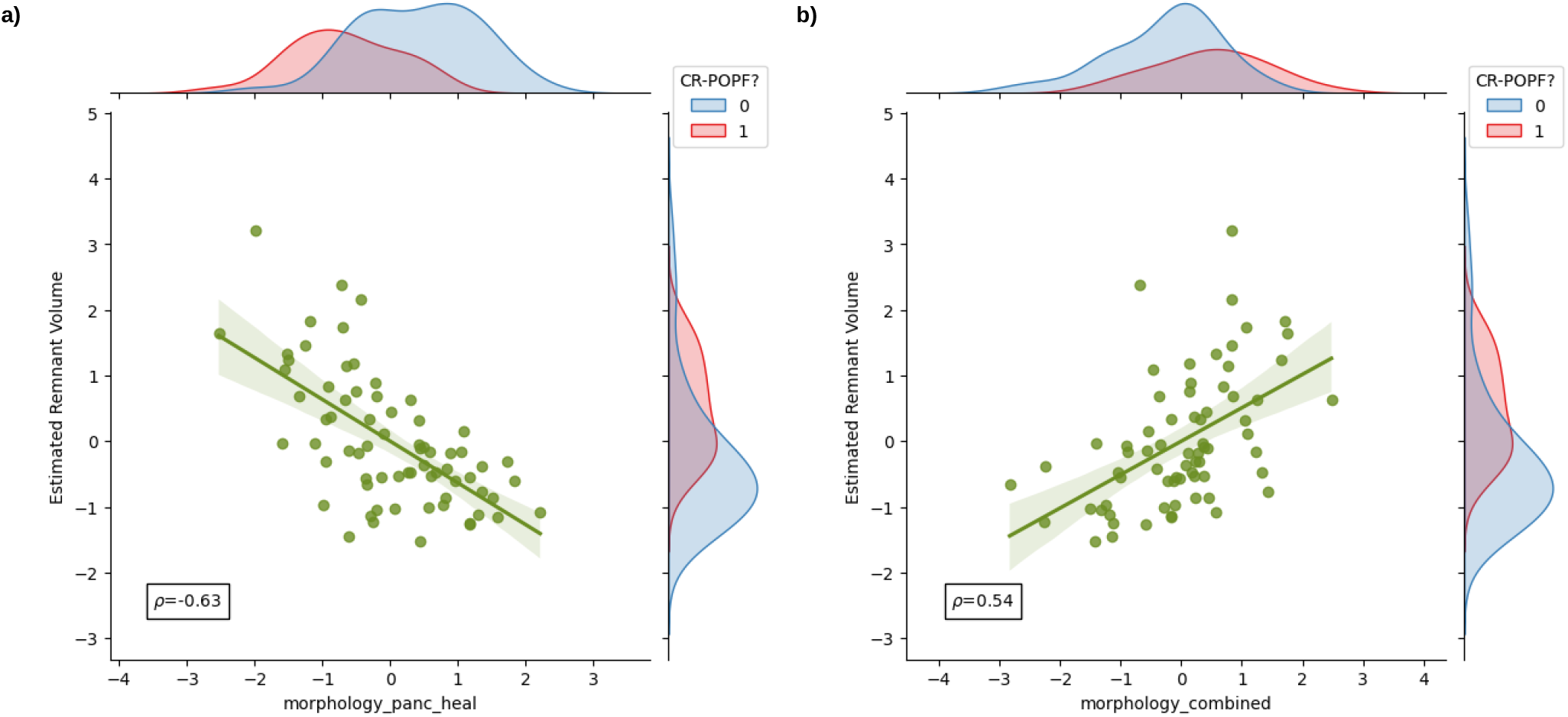
Comparison of radiomic features with standard CR-POPF risk factors. Comparison of estimated pancreatic remnant volume with - **a)**: the morphological feature *F*1_*M*_ of the healthy pancreatic segment; **b)**: the morphological feature *F*4_*M*_ of the combined segment. The marginal distributions for both axes are illustrated according to the CR-POPF outcome (0 – no CR-POPF, 1 - CR-POPF). The value *ρ* indicates the Spearman correlation co-efficient between the two variables. For comparability, the radiomic features and the estimated pancreatic remnant volume were standardised.

**Figure 4.**
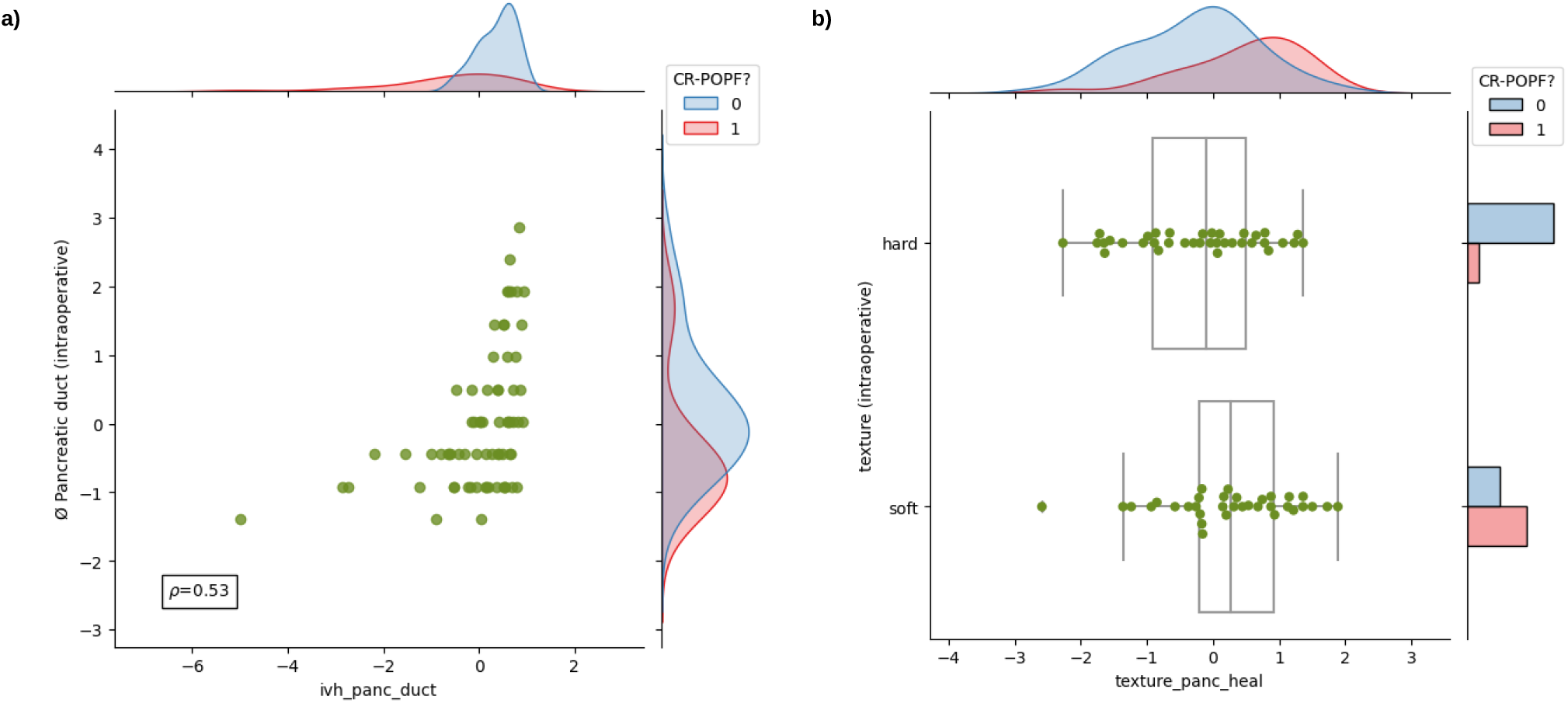
Comparison of radiomic features with standard CR-POPF risk factors. **a)**: Comparison of intraoperatively determined pancreatic duct width with the intensity volume histogram based feature *F*2_*IVH*_ of the pancreatic duct segment. The marginal distributions for both axes are illustrated according to the CR-POPF outcome (0 - no CR-POPF, 1 - CR-POPF). The value *ρ* indicates the Spearman correlation co-efficient between the two variables; **b)**: Comparison of intraoperatively determined pancreatic texture with the texture feature *F*3_*T*_ of the healthy pancreatic segment. For comparability, the radiomic features and the pancreatic duct width were standardised.

We also compared overall CR-POPF risk prediction performance of the combined radiomic and clinical signature with the stratification performance of the ua-FRS^13^, which is state-of-the-art for intraoperative CR-POPF risk stratification. The ua-FRS uses BMI, sex, intraoperative pancreatic texture and duct size to assess the risk of CR-POPF^13^. We identified a strong correlation between the predictions made by the combined radiomic and clinical signature based on preoperative data and the intraoperative ua-FRS (*ρ* = 0.7, Figure 5). Additionally, upon plotting confusion matrices for both methods, it was observed that the proposed preoperative signature was able to differentiate between the two classes better than the intraoperative ua-FRS (Supplementary Figure 5).

**Figure 5.**
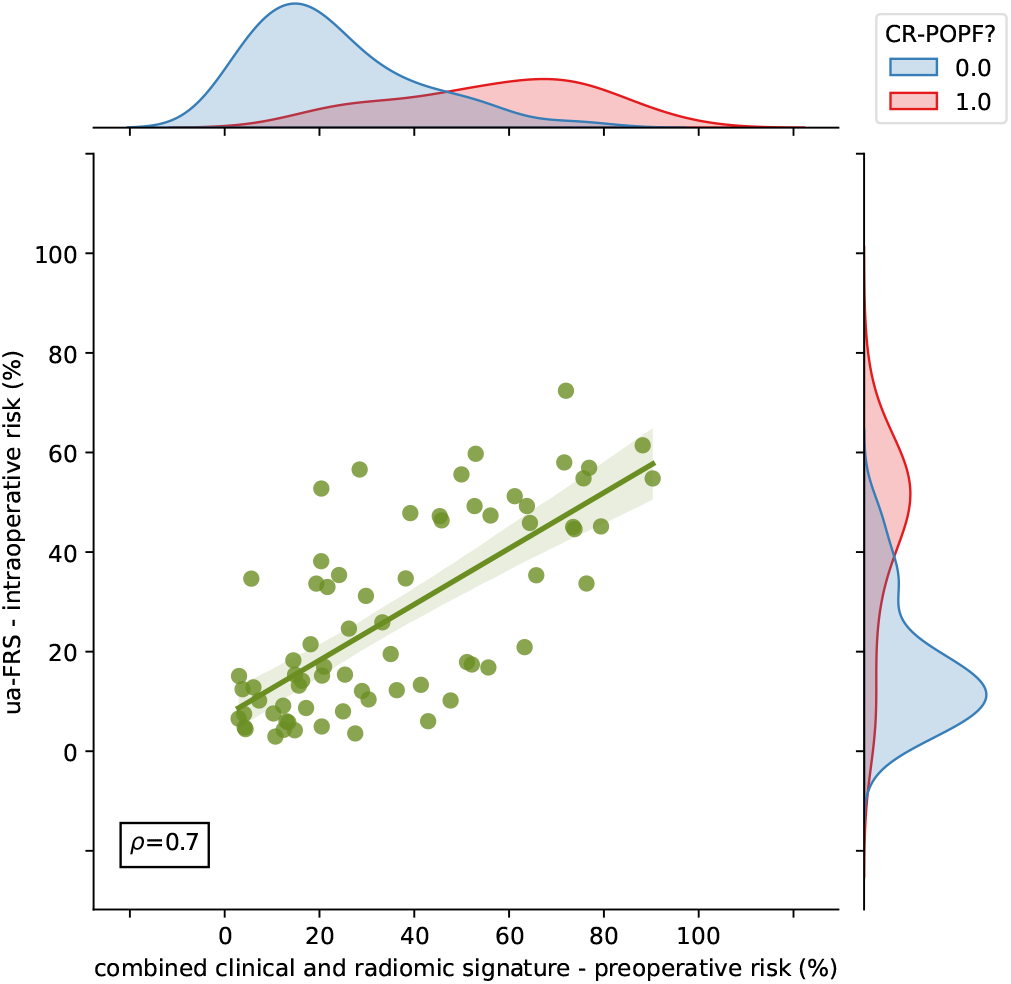
Comparison of CR-POPF risk predicted by the combined clinical and radiomic signature with the CR-POPF risk predicted by the intraoperatively determined ua-FRS (clinical gold standard). The marginal distributions for both axes are illustrated according to the CR-POPF outcome (0 - no CR-POPF, 1 - CR-POPF). The value *ρ* indicates the Spearman correlation co-efficient between the two variables.

These results indicate that the proposed combined radiomic and clinical signature developed solely based on preoperatively available clinical and routine imaging data can perform on par with the current state-of-the-art intraoperative models for CR-POPF risk stratification.

## Discussion

Affecting about 10-30% of patients undergoing pancreatic head resection^5–7^, CR-POPF is one of the most common complications after pancreatic head resection. In patients with pancreatic tumours (the most prevalent indication for pancreatic resection), a CR-POPF-related delay or suspension of indicated adjuvant therapy can drastically decrease patients’ chances for cure^3^. Therefore, accurate and early CR-POPF risk stratification has been intensively studied in the last decade^6,13^. The current gold standard of assessing CR-POPF risk is largely based on intraoperative factors such as parenchymal texture and probe-measured pancreatic duct diameter. However, an accurate and early risk assessment, ideally based on routinely available preoperative data, would facilitate risk-informed surgical planning and enable transparency in patient information^16^. To this end, we combined the available preoperative clinical data with annotated preoperative CT images to develop risk models for the prediction of CR-POPF in patients who underwent pancreatic head resection. The combined radiomic and clinical signature showed the best performance among the developed models with an AUC of 0.86 and a balanced accuracy score of 0.76 on the validation data. The proposed signature was developed using a combination of CT-derived radiomic features and clinical variables.

Limited interpretability of predictive machine learning tools has been linked with reduced trust in their performance and consequently, lower practical application^31,32^. Therefore, we explored associations between clinically validated CR-POPF risk factors and relevant radiomic features. On univariate analysis, the following features were found to be significantly associated with the outcome: (i) morphological (*F*1_*M*_) and (ii) texture (*F*3_*T*_) features of the healthy pancreatic segment; (iii) morphological feature (*F*4_*M*_) of the combined segment; (iv) intensity volume histogram based feature (*F*2_*IVH*_) of the pancreatic duct; and (v) BMI. The radiomic texture feature (*F*3_*T*_) captures busyness or large changes in the discretised grey levels between neighbouring voxels^21^. The value of *F*2_*IVH*_ represents the difference between the largest volume fractions (*V*_10_ and *V*_90_) containing intensity fractions of at least 10% and 90%^21^ in the pancreatic duct segment. Pancreatic texture, pancreatic duct width, and BMI are established CR-POPF risk factors^6,13,16,33^.

The radiomic feature *F*1_*M*_ describes the surface to volume ratio of the healthy pancreatic segment. The morphological feature *F*4_*M*_ measures the extent of the axis along which the variation of data is maximum in the combined segment. The feature expression map (Figure 2) revealed an inverse relationship between *F*1_*M*_ and the predicted risk of CR-POPF, and a direct relationship between *F*4_*M*_ and the predicted risk of CR-POPF. A similar trend was observed between the features *F*1_*M*_ and *F*4_*M*_, and the estimated PRV (Figure 3). Therefore, the results of the proposed signature are in line with previous studies^16,17^ that indicate an increased risk of CR-POPF for patients with higher estimated PRV. Moreover, the correlation between the intensity volume histogram based feature *F*2_*IVH*_ of the pancreatic duct segment with the intraoperatively assessed pancreatic duct width and between the texture feature *F*3_*T*_ of the healthy pancreatic segment and intraoperatively determined pancreatic texture could explain the relevance of these features and imply common relationship with the overall CR-POPF risk.

Ultimately, the observed association (Spearman correlation coefficient, *ρ* = 0.7) between the intraoperative ua-FRS^13^ and the predictions made by the proposed preoperative signature underlines its relevance and validity (Figure 5).

The results of this study suggest that radiomic features from preoperative CT imaging and clinical factors can help identify patients at high risk for postoperative pancreatic fistula following pancreatic head resection. However, the practical implications of preoperative risk stratification remain to be clarified. In general, management options in patients with a known high risk of CR-POPF include different anastomotic techniques^34,35^, application of somatostatins^36,37^, adapted drain management^38,39^ as well as intensified follow-up^40^ and changes in the overall surgical approach. In this context, recent studies provide evidence that in selected patients, primary total pancreatectomy, despite being associated with considerable postoperative morbidity^41,42^, could provide benefits over pancreatic head resection with a high-risk pancreatic anastomosis^43–46^. This option could be particularly viable in patients with preoperatively impaired glucose tolerance or when concomitant islet cell autotransplantation is an option^47,48^.

The limitations of this work are mostly related to the monocentric and retrospective nature of design. The consequent limited availability of data led to the combined radiomic and clinical signature being developed over the same patient cohort that was used to select the features for the radiomic signature. Consequently, the results of the signature cannot be considered as validation results in a true sense. Therefore, an external validation of the signature will be carried out in future. Moreover, some of the selected radiomic features were strongly correlated with the mesh-based volumes of their corresponding anatomical structures (Supplementary Figure 4), which hindered the performance of the risk model developed using a combination of these features (Supplementary table 7). On the whole, there is potential for the improvement of feature selection strategy in future. In the analysed cohort, 33 out of 108 patients developed CR-POPF. The resulting CR-POPF rate of 31% is higher than CR-POPF rates previously reported in the literature^5,6,49^. These shortcomings are likely due to selection bias related to the retrospective nature of the study design. In addition, patients with clinical symptoms are interventionally treated relatively early based on institutional standards, which could result in a trend towards overestimation of the proportion of patients with grade B POPF. On the CT level, patients with comorbidities and imaging findings that would substantially impact image quality (i.e. calcifying chronic pancreatitis or a stent in the common bile duct) were excluded from this study. We also observed substantial variation in the time interval between CT imaging and pancreatic head resection. We observed negligible influence (Spearman correlation coefficient, *ρ* = 0.03) of this delay on the CR-POPF risk predicted by the proposed preoperative signature. However, a large scale analysis of the impact of such observed time delay and imaging findings on the predictive power of the proposed signature is imminent. To overcome these limitations and validate the proposed signature, a respective study on a well balanced and sufficiently large multicentric patient cohort is in preparation.

In summary, the results of this study indicate that preoperative data like CT-based radiomic features and relevant clinical variables can contribute to accurate preoperative CR-POPF risk assessment in patients undergoing pancreatic head resection. An integration of the proposed risk assessment tool in clinical workflows could aid the surgical decision making process and ultimately contribute to improved patient outcomes in future.

## Data Availability

All data produced in the present study are available upon reasonable request to the authors.

## Data Availability

The datasets generated and analysed during the current study are available from the corresponding author on reasonable request.

## Acknowledgements

The research was carried out within the Surgomics project funded by the German Federal Ministry of Health, University Hospital Heidelberg and Uniklinikum Carl Gustav Carus, Dresden. Partners from industry: Karl Storz GmbH and Phellow Seven GmbH.

## Author contributions statement

Stefan Leger (S.L.), Fiona R. Kolbinger (F.R.K.) and Nithya Bhasker (N.B.) conceived the study; S.L., F.R.K., N.B., Alexander Zwanenburg (A.Z.) and Steffen Löck (S. Löck) designed the experiments; A.Z., S.L. and N.B. developed the software; S.L., F.R.K. and N.B. conducted the experiments; N.B. and S.L. visualised the results; S.L., F.R.K., N.B., A.Z. and S. Löck analysed the results; Stefanie Speidel (S.S.), S. Löck, Jürgen Weitz (J.W.), Jens-Peter Kühn (J.K.), Marius Distler (M.D.), and Ralf-Thorsten Hoffmann (R.H.) provided resources and supervision; F.R.K., Nadiia Skorobohach (N.S.), J.K. and M.D. curated the clinical data; N.B., F.R.K., S.L. and S.S. drafted the manuscript; N.B., F.R.K., N.S., S.S., S.L., A.Z., S.Löck, J.W., R.H., M.D. and J.K. reviewed the manuscript draft; All authors read and agreed to the final version of the manuscript.

## Ethics declarations

Ethical approval for the retrospective analyses of clinical and imaging data was obtained from the Ethics Committee at the Technische Universität Dresden (BO-EK-263062020). All analyses were carried out in accordance with the relevant guidelines and regulations, in particular the Declaration of Helsinki and its later amendments. The local institutional review board (Ethics Committee at the Technische Universität Dresden) approved the study protocol (BO-EK-263062020) and waived written patient consent for the retrospective analysis of routinely acquired anonymised clinical data.

## Additional information

### Supplementary information

Refer to the Supplementary information online for more details about the study and additional results.

