## Supplementary Data for "Prediction of clinically relevant postoperative pancreatic fistula using radiomic features and preoperative data"

[1]National Center for Tumor Diseases (NCT/UCC) Partner Site Dresden, Germany
[2]Department of Visceral, Thoracic and Vascular Surgery, University Hospital Carl Gustav Carus, Technische Universität Dresden, Dresden, Germany
[3]Else Kröner Fresenius Center for Digital Health, TU Dresden, Dresden, Germany
[4]Institute and Polyclinic for Diagnostic and Interventional Radiology, University Hospital Carl Gustav Carus Dresden, TU Dresden, Dresden, Germany
[5]OncoRay - National Center for Radiation Research in Oncology, Faculty of Medicine and University Hospital Carl Gustav Carus, Technische Universität Dresden, Helmholtz-Zentrum Dresden-Rossendorf, Dresden, Germany
[*]
[+]these authors contributed equally to this work
[++]shared last authorship

#### ABSTRACT

Clinically relevant postoperative pancreatic fistula (CR-POPF) can significantly affect the treatment course and outcome in pancreatic cancer patients. Preoperative prediction of CR-POPF can aid the surgical decision-making process and lead to better perioperative management of patients. In this retrospective study of 108 pancreatic head resection patients, we present risk models for the prediction of CR-POPF that use combinations of preoperative computed tomography (CT)-based radiomic features, mesh-based volumes of annotated intra- and peripancreatic structures and preoperative clinical data. The risk signatures were evaluated and analysed in detail by visualising feature expression maps and by comparing significant features to the established CR-POPF risk measures. Out of the risk models that were developed in this study, the combined radiomic and clinical signature performed best with an average area under receiver operating characteristic curve (AUC) of 0.86 and a balanced accuracy score of 0.76 on validation data. The following pre-operative features showed significant correlation with outcome in this signature ($p < 0.05$)- texture and morphology of the healthy pancreatic segment, intensity volume histogram-based feature of the pancreatic duct segment, morphology of the combined segment, and BMI. The predictions of this pre-operative signature showed strong correlation (Spearman correlation co-efficient, $\rho = 0.7$) with the intraoperative updated alternative fistula risk score (ua-FRS), which is the clinical gold standard for intraoperative CR-POPF risk stratification. These results indicate that the proposed combined radiomic and clinical signature developed solely based on preoperatively available clinical and routine imaging data can perform on par with the current state-of-the-art intraoperative models for CR-POPF risk stratification.

### 1 Detailed patient characteristics

| Variable | Statistic | CR-POPF | Non CR-POPF |
|---|---|---|---|
| Patients | n | 33 | 75 |
| Pre-existing diabetes: | n | | |
| yes | | 8 | 20 |
| no | | 25 | 55 |
| History of smoking: | n | | |
| yes | | 4 | 15 |
| no | | 29 | 60 |
| History of alcohol abuse: | n | | |
| yes | | 4 | 13 |
| no | | 29 | 62 |
| CA 19-9: | n | | |
| Normal (< 24 U/mL) | | 11 | 13 |
| Abnormal (> 24 U/mL) | | 22 | 62 |
| Preoperative amylase: | n | | |
| Normal (< 0.88 $\mu$mol/s*L) | | 26 | 61 |
| Abnormal (> 0.88 $\mu$mol/s*L) | | 7 | 14 |
| Preoperative lipase: | n | | |
| Normal (< 1.0 $\mu$mol/s*L) | | 20 | 52 |
| Abnormal (> 1.0 $\mu$mol/s*L) | | 13 | 23 |
| Mesh-based volumes of segmented structures ($cm^3$): | mean $\pm$ standard deviation | | |
| pancreatic duct | | 2.71 $\pm$ 2.38 | 5.21 $\pm$ 4.62 |
| healthy pancreas | | 95.80 $\pm$ 43.65 | 62.81 $\pm$ 30.93 |
| pancreatic pathology | | 18.15 $\pm$ 28.41 | 15.27 $\pm$ 15.53 |
| arteries | | 55.91 $\pm$ 24.96 | 45.50 $\pm$ 15.40 |
| bile duct | | 15.21 $\pm$ 21.58 | 13.36 $\pm$ 13.55 |
| portal vein | | 36.68 $\pm$ 14.58 | 33.92 $\pm$ 11.00 |

**Supplementary Table 1.** Detailed patient characteristics of the retrospective cohort used in the study.

#### 2 Configuration settings for radiomic feature extraction

| Parameters | Values |
|---|---|
| Image interpolation method | linear |
| Image voxel size | $1 \times 1 \times 1\text{mm}^3$ |
| ROI interpolation method | linear |
| Partial volume ROI inclusion threshold | 0.5 |
| Discretisation method | fixed bin number, $n = 32$ |
| Intensity volume histogram discretisation method | fixed bin number, $n = 1000$ |
| Texture matrices computed | Grey-level co-occurrence matrix (GLCM), Grey-level run length matrix (GLRLM), Grey-level size zone matrix (GLSZM), Grey-level distance zone matrix (GLDZM), Neighbourhood grey tone difference matrix (NGTDM), Neighbourhood grey level dependence matrix (NGDM) |
| Calculation method for texture matrices | 3D |
| Merge method for GLCM and GLRLM | average |
| GLCM distance (in voxels) for determining neighbourhood | 1, 2, 3 |
| NGLDM distance (in voxels) for determining neighbourhood | 1.8 |
| NGLDM difference level (alpha) | 0 |
| Spatial filters for image transformation | wavelet (coiflet-1), Laplacian of Gaussian (LoG) |
| LoG filter width, $\sigma$ (mm) | 5, 4, 3, 2, 1, 0.5 |

**Supplementary Table 2.** Parameters used for the extraction of radiomic features.

#### 3 Selection of features for radiomic signature

A set of preliminary risk models were developed by using the computed radiomic features of 148 patients who underwent pancreatic surgery. These patients included patients who underwent pancreatic head resection ($n = 108$) and other surgeries ($n = 40$). The radiomic features were extracted from anatomical structures in the preoperative CT data and therefore, the radiomic feature extraction process was agnostic to the kind of surgery or the postoperative CR-POPF outcome. Moreover, upon conducting a Chi-squared test, we found that the distributions of outcome in head and non-head resection patient groups in our cohort showed no significant difference ($p = 0.42$). Table 3 summarises the results of the statistical analysis and Table 4 compares the performance of the preliminary radiomic risk model across different patient groups.

| | Observed (O) | | Expected(E) | |
|---|---|---|---|---|
| | Head resection | Other surgeries | Head resection | Other surgeries |
| CR-POPF | 33 | 15 | 35.03 | 12.97 |
| No CR-POPF | 75 | 25 | 72.97 | 27.03 |

**Supplementary Table 3.** Summary of Chi-squared test conducted to compare the distribution of CR-POPF outcome in head resection and non-head resection patient groups. Null hypothesis: The distribution of CR-POPF outcome is similar between the two groups. Chi-squared test value, $\chi^2 = 0.64$; Degrees of freedom, dof = 1; p-value = 0.42. Null hypothesis cannot be rejected. The differences in the distributions of CR-POPF outcome between the two groups is not statistically significant for the tested cohort.

Subsequently, representative features for patients who underwent pancreatic head resection ($n = 108$) were selected based on the permutation feature importance method. That is, features were deemed important and selected based on the reduction in model performance over the head resection patients when the most important features (based on feature occurrence) of the preliminary risk models were permuted. Due care was observed in this process, to not choose features that were strongly correlated (absolute Spearman correlation coefficient, $\rho > 0.75$) with each other. The features selected for the radiomic signature are described in Table 5.

| | Exploratory data | | Validation data | |
|---|---|---|---|---|
| **Patient group** | **AUC** | **Bal. acc.** | **AUC** | **Bal. acc.** |
| All pancreatic surgeries ($n = 148$) | $0.91 \pm 0.05$ | $0.68 \pm 0.09$ | $0.70 \pm 0.03$ | $0.55 \pm 0.03$ |
| Head resection ($n = 108$) | $0.93 \pm 0.04$ | $0.70 \pm 0.09$ | $0.74 \pm 0.04$ | $0.56 \pm 0.04$ |
| Other surgeries ($n = 40$) | $0.85 \pm 0.11$ | $0.65 \pm 0.11$ | $0.54 \pm 0.02$ | $0.51 \pm 0.02$ |

**Supplementary Table 4.** Mean of average area under the receiver operating characteristic curve (AUC) and balanced accuracy score (Bal. acc.), and their standard deviations for preliminary risk models over all combinations of feature selection methods and machine learning algorithms for the different patient groups.

| Feature | Description | Segment | Image |
|---|---|---|---|
| $F1_M$ | morphological feature - surface to volume ratio | panc_heal | base |
| $F2_{IVH}$ | volume fraction difference between intensity fractions (diff_v10_v90) | panc_duct | wavelet coiflet-1 (hhl) |
| $F3_T$ | NGTDM feature - busyness | panc_heal | LoG |
| $F4_M$ | morphological feature - major axis length | combined | base |

**Supplementary Table 5.** Description of features selected for radiomic signature.

#### 4 Performance of the risk models

Table 6 gives an overview of the risk models developed with different input feature combinations. The performance of the models was assessed using average area under the receiver operating characteristic curve (AUC) and balanced accuracy score on the ensemble predictions. The heatmaps representing the performances of clinical and volume model over different combinations of feature selection (FS) and machine learning (ML) methods are illustrated in Figures 1 and 2. To ensure robustness, one combination of FS and ML method was selected as the best based on the following scheme[1]: (i) median performance of each FS method over all ML algorithms and vice versa was determined on the exploratory data; (ii) the FS and ML methods with AUCs closest to the respective median performances were chosen as the best combination of methods. The AUC and balanced accuracy score of the different risk models for these best FS and ML method are tabulated in Table 7.

| Model | Selected radiomic features | Clinical variables | Volumes of segments | Feature selection applied |
|---|---|---|---|---|
| clinical | ✗ | ✓ | ✗ | ✓ |
| volume | ✗ | ✗ | ✓ | ✓ |
| radiomic signature | ✓ | ✗ | ✗ | ✗ |
| combined radiomic and clinical signature | ✓ | ✓ | ✗ | ✗ |
| combined radiomic and volume signature | ✓ | ✗ | ✓ | ✗ |
| combined radiomic, clinical and volume signature | ✓ | ✓ | ✓ | ✗ |

**Supplementary Table 6.** Overview of developed risk models with the combinations of input features used.

The features having a significant association with the CR-POPF outcome ($p < 0.05$) were standardised, and heatmaps of the resulting standardised features were visualised for the entire patient cohort in order to qualitatively assess the trends and correlations with the outcome. Figures 3 illustrates such a feature expression map for the combined radiomic, clinical and volume signature.

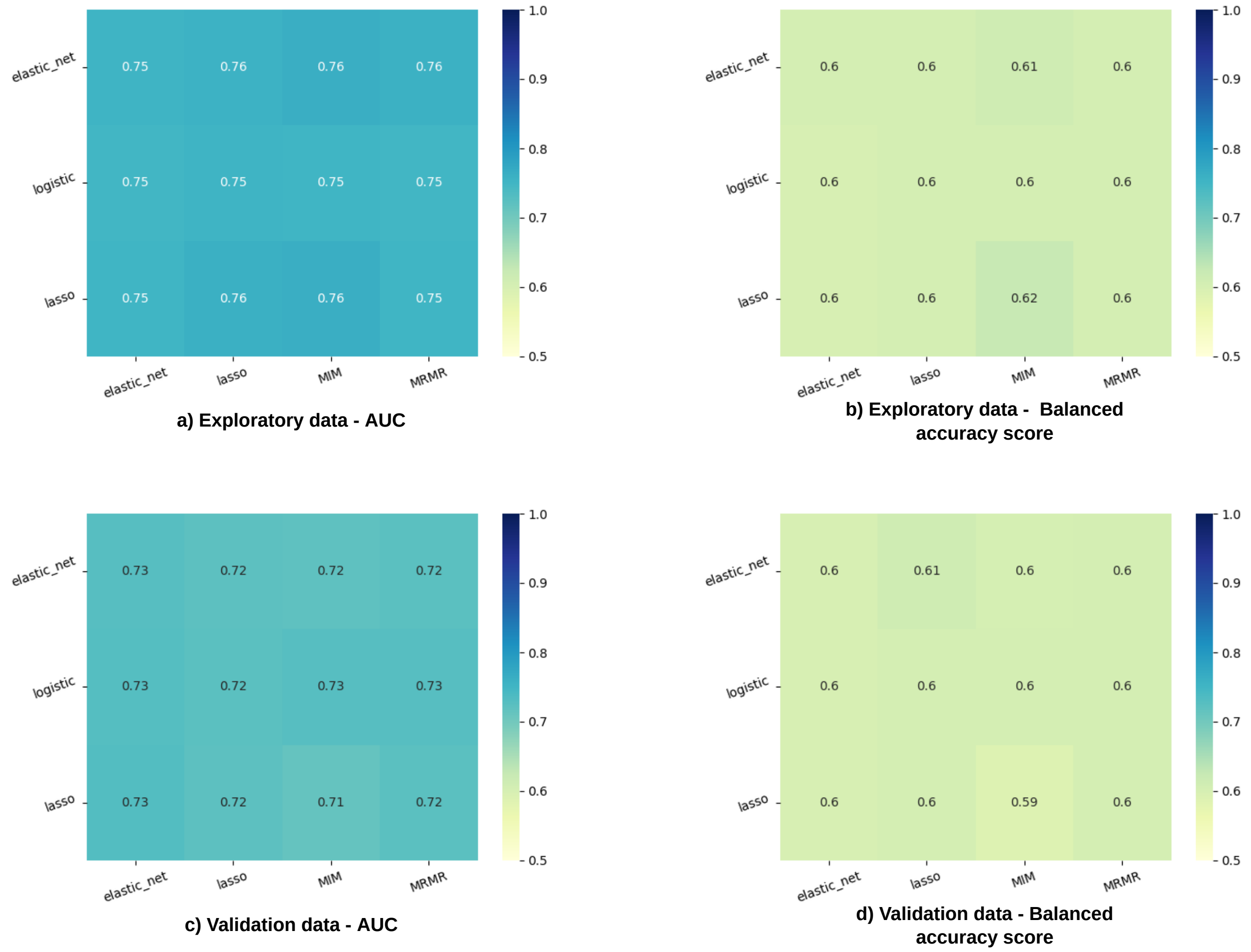


**Supplementary Figure 1.** Heatmaps representing the performance of different feature selection (columns) and learning method (rows) combinations for the exploratory and validation data of the clinical model.

| Model | Exploratory data | | Validation data | | best FS | best ML |
|---|---|---|---|---|---|---|
| | AUC | Bal. acc. | AUC | Bal. acc. | | |
| clinical | 0.75 | 0.60 | 0.73 | 0.60 | elastic-net | elastic-net |
| volume | 0.85 | 0.74 | 0.83 | 0.72 | lasso | elastic-net |
| radiomic signature | 0.87 | 0.71 | 0.85 | 0.71 | - | elastic-net |
| combined radiomic and clinical signature | 0.90 | 0.79 | 0.86 | 0.76 | - | elastic-net |
| combined radiomic and volume signature | 0.88 | 0.72 | 0.84 | 0.69 | - | elastic-net |
| combined radiomic, clinical and volume signature | 0.91 | 0.80 | 0.86 | 0.71 | - | elastic-net |

**Supplementary Table 7.** Average area under the receiver operating characteristic curve (AUC) and balanced accuracy score (Bal. acc.) for the best combination of feature selection methods and machine learning algorithms. Abbreviations: FS - feature selection, ML - machine learning, elastic-net - logistic regression with elastic-net penalty, lasso - logistic regression with lasso penalty.

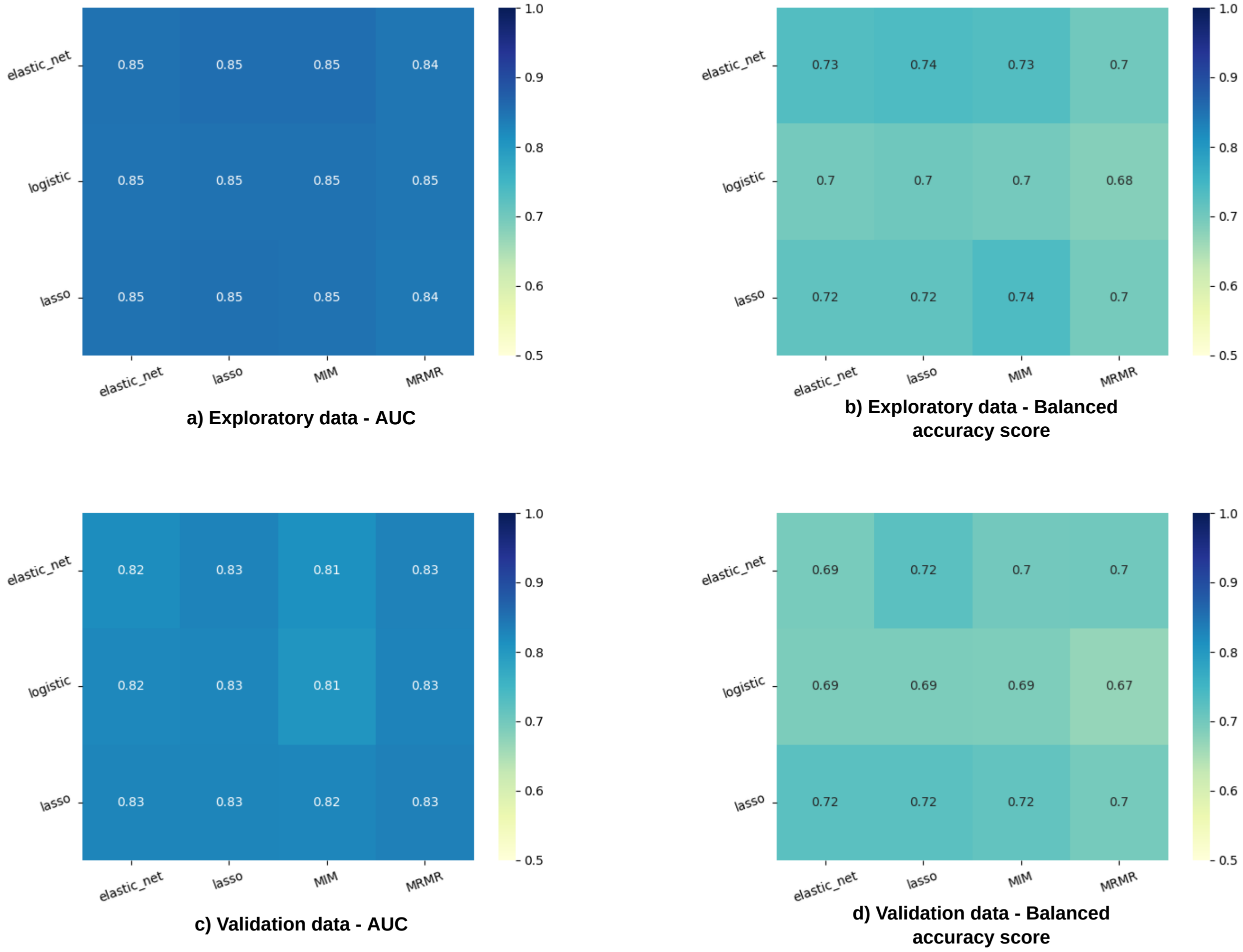


**Supplementary Figure 2.** Heatmaps representing the performance of different feature selection (columns) and learning method (rows) combinations for the exploratory and validation data of the volume model.

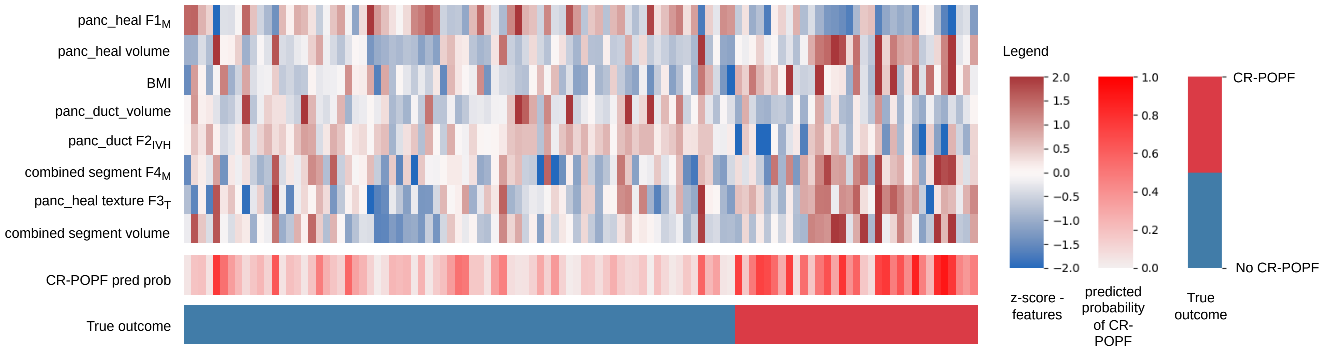


**Supplementary Figure 3.** Feature expression map for the combined radiomic, clinical and volume signature. The values indicated in the heatmaps are z-scores of the standardised features, probability of developing CR-POPF as predicted by the model and the true outcome for all the patients included in the study. Abbreviations: pred prob – prediction probability, $F_M$ – morphological feature, $F_T$ – texture feature, $F_{IVH}$ - intensity volume histogram based feature.

#### 5 Feature associations

The features selected for radiomic signature showed considerable association with the mesh-based volumes of the corresponding anatomical structures. Figure 4 illustrates the relationship between these features.

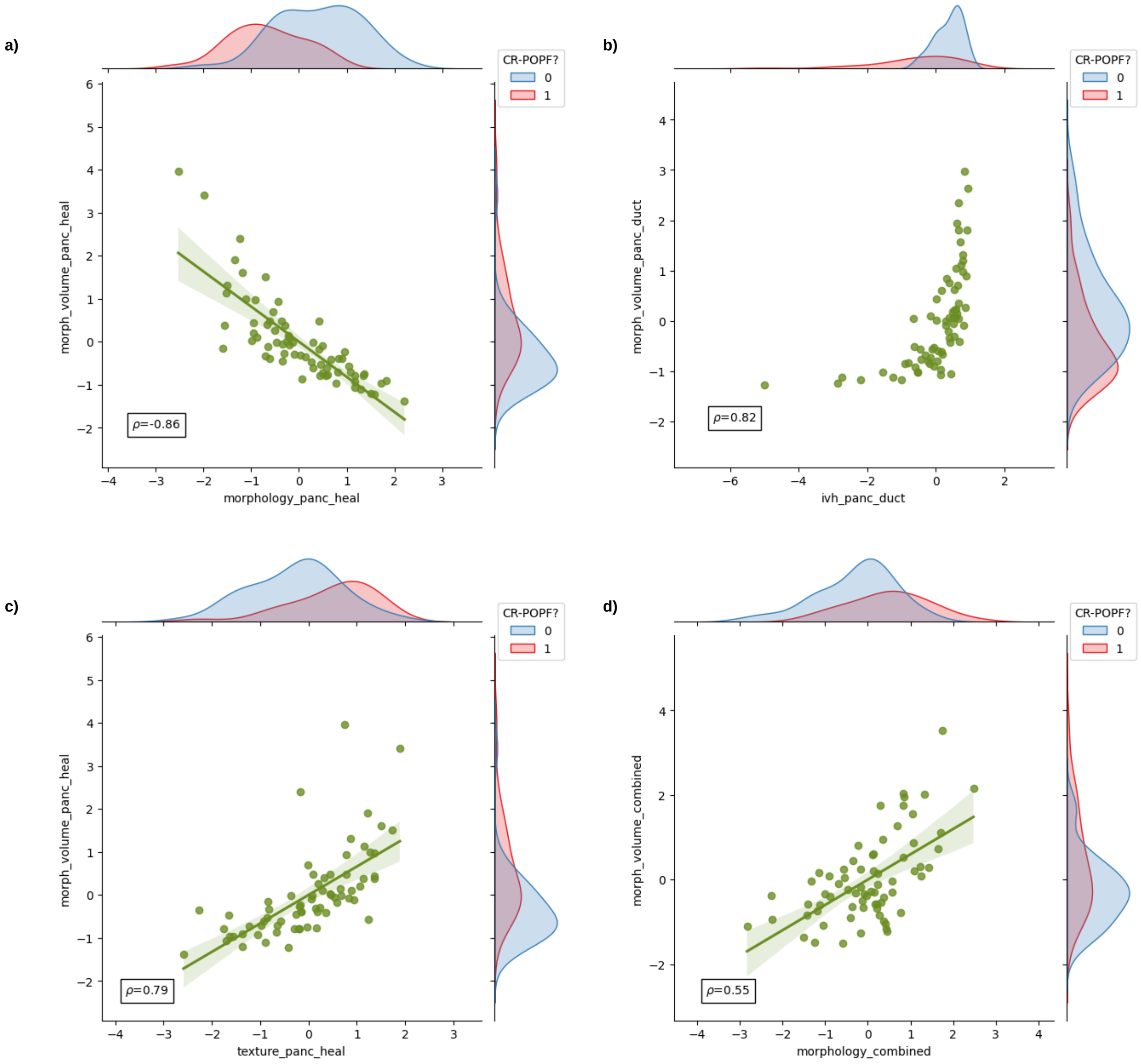


**Supplementary Figure 4.** Comparison of selected radiomic features with mesh-based volumes of the corresponding segments. The marginal distributions for both axes categorised according to the CR-POPF outcome (0 - no CR-POPF, 1 - CR-POPF) are also illustrated. The value $\rho$ indicates the Spearman correlation co-efficient between the two variables.

#### 6 Comparison with ua-FRS

The intraoperative risk measures were available for 70 out of the 108 patients considered for this study. A threshold of 20% was considered for the classification of CR-POPF based on ua-FRS[2]. Figure 5 illustrates the comparison between the confusion matrices of intraoperative ua-FRS and our preoperative combined radiomic and clinical signature.

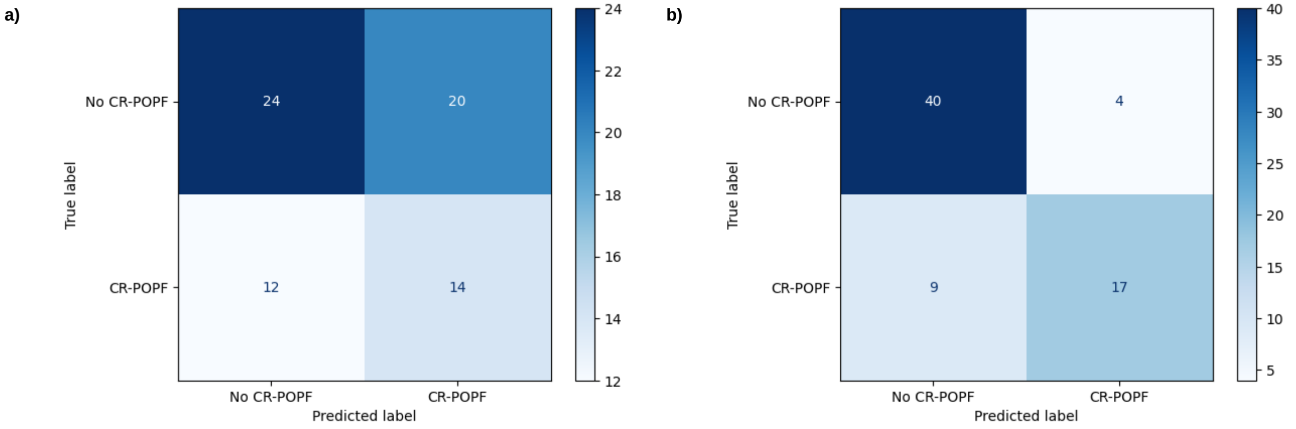


**Supplementary Figure 5.** Comparison of confusion matrices of a) intraoperative ua-FRS with b) our preoperative combined radiomic and clinical signature.
